# Hyperglycemic exacerbation of myocardial infarction through SGLT1 - a glucose paradox

**DOI:** 10.1101/2023.03.07.23286959

**Authors:** Alhanoof Almalki, Sapna Arjun, Idris Harding, Hussain Jasem, Maria Kolatsi-Joannou, Daniyal J Jafree, David A Long, Derek M Yellon, Robert M Bell

**Affiliations:** Hatter Cardiovascular Institute, Institute for Cardiovascular Science, University College London, London, UK; Developmental Biology and Cancer Programme, UCL Great Ormond Street Institute of Child Health, London, UK; UCL MB/PhD Programme, Faculty of Medical Sciences, University College London, London, UK

**Keywords:** SGLT1, SGLT2, myocardial infarction, reperfusion injury, diabetes, hyperglycemia

## Abstract

**Background:** Hyperglycemia is common during acute myocardial infarction, irrespective of diabetic status, and portends excess mortality. The mechanisms of this adverse outcome remain unelucidated.

**Objectives:** To test the hypothesis that elevated glucose, at the time of reperfusion following myocardial ischemia, is directly injurious to the heart through induction of sodium/glucose-linked transporter 1 (SGLT1) activity.

**Methods:** *Ex-vivo*, Langendorff rodent models of 35minute global ischemia and 2hour reperfusion injury were utilised, with variable glucose and reciprocal mannitol concentrations maintaining equivalent osmolarity across groups during reperfusion. Infarct size was assessed by tri-phenyltetrazolium staining. SGLT1 expression was determined in rodents by rtPCR, RNAscope and immunohistochemistry and in human with single-cell transcriptomic analysis. *Ex-vivo*, functional involvement of SGLT1 was determined using three, structurally distinct pharmacological inhibitors: phlorizin, canagliflozin and mizagliflozin.

**Results:** In non-diabetic rodent hearts there was a J-shaped dose-response relationship between reperfusion-glucose concentration and infarct size, an association ameliorated in diabetic heart. Single-cell transcriptomic analysis revealed human myocardial SGLT1 expression equivalent to that seen in rodents at both an RNA and protein level. Diabetic rodent heart SGLT1 expression was significantly reduced compared to non-diabetic, and pharmacological SGLT1 inhibition abrogated excess injury associated with high glucose in non-diabetic heart.

**Conclusion:** Elevated glucose during reperfusion exacerbated myocardial infarction in non-diabetic heart, but this exacerbation was attenuated in diabetic rat heart where SGLT1 expression is suppressed. Inhibiting non-diabetic heart SGLT1 abrogates the excess injury associated with elevated glucose, thus highlighting SGLT1 as a potential clinical translational target to improve outcomes in acute myocardial infarction associated with hyperglycemia.

## Background

Despite improvements in the management of acute myocardial infarction (AMI), ischemic heart disease remains the leading cause of death worldwide^1^. Among patients with an acute coronary syndrome (ACS), 25-50% will have hyperglycemia at the time of presentation, irrespective of the presence or absence of diabetes mellitus (DM)^2^ and is recognised as an adverse marker for cardiovascular morbidity^3^ and mortality^2^. Epidemiological studies reveal a linear relationship between increasing blood glucose and excess death – a relationship more pronounced in patients without a prior diagnosis of DM^2^. However, demonstration of a causal relationship between glucose and cardiovascular mortality in ACS has proven challenging. The Diabetes Mellitus Insulin Glucose Infusion in Acute Myocardial Infarction^4^, a trial of intense glucose control in ACS patients with a glucose greater than 11mmol/L, irrespective of diabetic status, resulted in significant improvements in cardiovascular outcomes. This data suggested that managing hyperglycemia would be beneficial, however, subsequent studies^5,6^ failed to reproduce the earlier study’s reduction of cardiovascular mortality. Thus, it remains unclear whether elevated glucose during AMI is directly injurious or glucose is simply a biomarker of poor cardiovascular outcome.

While the mechanistic connection between glucose and ACS outcomes remains controversial, the adverse association between glucose and outcomes are recognised in national and international guidelines, with recommendations to control hyperglycemia^7,8^. However, these guidelines also recognise that aggressive glycemic control is potentially hazardous: hypoglycemia also drives increased mortality^7,8^. The relationship between blood glucose and mortality is complex, with a dose-response relationship following a “J-curve”: the lowest mortality coinciding with euglycemia in non-diabetic patients (around 5.5mmol/L), with upward inflections of the mortality curve with both hypo- and hyperglycemia^2^.

Sodium/glucose linked transport (SGLT) has emerged as a target for improving outcomes in patients with heart failure where inhibition of the SGLT2 member of the SLC5A solute carrier family reduces hospitalisations and improves cardiovascular outcomes^9^. However, SGLT2 is expressed predominantly in the kidney proximal tubule with lower expression in other tissues and absent in the heart^10,11^. Thus, the cardiovascular benefits from SGLT2 inhibition are indirect, and the mechanisms of action unknown. The other significant glucose transporter in the SLC5A family is SGLT1. It is expressed within the kidney, but also plays a role in glucose absorption from the gut and is expressed in the heart^12^, although its physiological and pathophysiological roles in the myocardium are unclear.

Myocardial SGLT1 expression is upregulated in models of left ventricular dysfunction, hypertrophy and following AMI^13^. Diabetes also alters SGLT1 expression, but reports are mixed as to whether DM up- or down-regulates SGLT1^14^. SGLT1 activity is glucose-dependent, and unlike SGLT2 that transports sodium and glucose at a ratio of 1:1, the SGLT1 sodium:glucose ratio is 2:1. Thus, glucose-driven SGLT1 activity leads to greater sodium influx than SGLT2 under similar conditions. With acute ischemic stress, increased intracellular sodium loading of the cell driven by elevated extra-cellular glucose may imperil viability and survival. This, together with the variances of SGLT1 expression with disease, makes SGLT1 a potential target for understanding the excess injury associated with elevated glucose at the time of AMI.

In this study, we wanted to address three questions: (1) is there is a pathophysiological link between glucose at the time of reperfusion and infarct size; (2) is SGLT1 activity, in whole or in part, responsible for excess myocardial injury, and (3) does DM alter SGLT1 expression and does this correlate to the apparent increased resistance to hyperglycemia in diabetics.

## Methods

### Animals

Animal use complied with Animals (Scientific Procedures) Act 1986 Amendment Regulations 2012 and was approved by UCL AWERB and UK Home Office. Sprague-Dawley (SD) rats (300-390g; UCL Biological Services), Zucker Diabetic Fatty rats (ZDF) and Zucker lean nondiabetic rats (ZL) (8-10 weeks; Charles River Laboratories) were used. ZDF rats were fed Purina #5008 diet to induce Type 2 diabetes. SD rats and ZL rats were maintained on Purina #5001.

### Reagents

SGLT inhibitors: Canagliflozin (Janssen R&D) and Mizagliflozin (MedChem Express LLC). Cardiomyocyte isolation/culture reagents: Protease XIV and Laminin (Sigma-Aldrich), Collagenase Type 5 (Lorne Labs, UK). Medium 199 and routine biochemicals (Thermofisher Scientific). RNA isolation and PCR kit (Qiagen) and RNAscope® Multiplex Fluorescent Detection Kit (ACD, Biotechne).

### *Ex-vivo* cardiac ischemia-reperfusion

*Ex-vivo* perfusion of rodent hearts were carried out as described^15^. Briefly, animals were anesthetised using pentobarbital sodium (100-150mg/kg) and heparin sodium (400-500u/kg) IP. Hearts were harvested and retrogradely perfused with modified Krebs-Henseleit buffer containing 118 mmol/L NaCl, 25mmol/L NaHCO_3_, 11mmol/L D-glucose, 4.7mmol/L KCl, 1.22mmol/L MgSO_4_·7H_2_O, 1.21mmol/L KH_2_PO_4_ and 1.84mmol/L CaCl_2_.2H_2_O (pH 7.4, 37°C, perfusion pressure 70-80mmHg). Following 20min of stabilisation perfusion, hearts were subjected to global ischemia for 35minutes followed by 2-hour reperfusion. Hearts were randomized to receive reperfusion buffer containing variable glucose concentrations from 5.5 to 22mmol/L glucose with reciprocal concentration of mannitol (16.5 to 0mmol/L) to maintain osmolarity (**Fig.1A**). In SGLT inhibitor experiments, hearts were randomized to receive either vehicle (DMSO) or drug (3μmol/L phlorizin, 5nmol/L or 1μmol/L Canagliflozin, or 100nM Mizagliflozin), alongside either 11mmol/L glucose + 11mmol/L mannitol or 22mmol/L glucose. After reperfusion, heart infarct size (% of LV area) was determined using the viability stain 2,3,5-triphenyltetrazolium chloride followed by blinded analysis using Image J (NIH).

**Figure 1:**
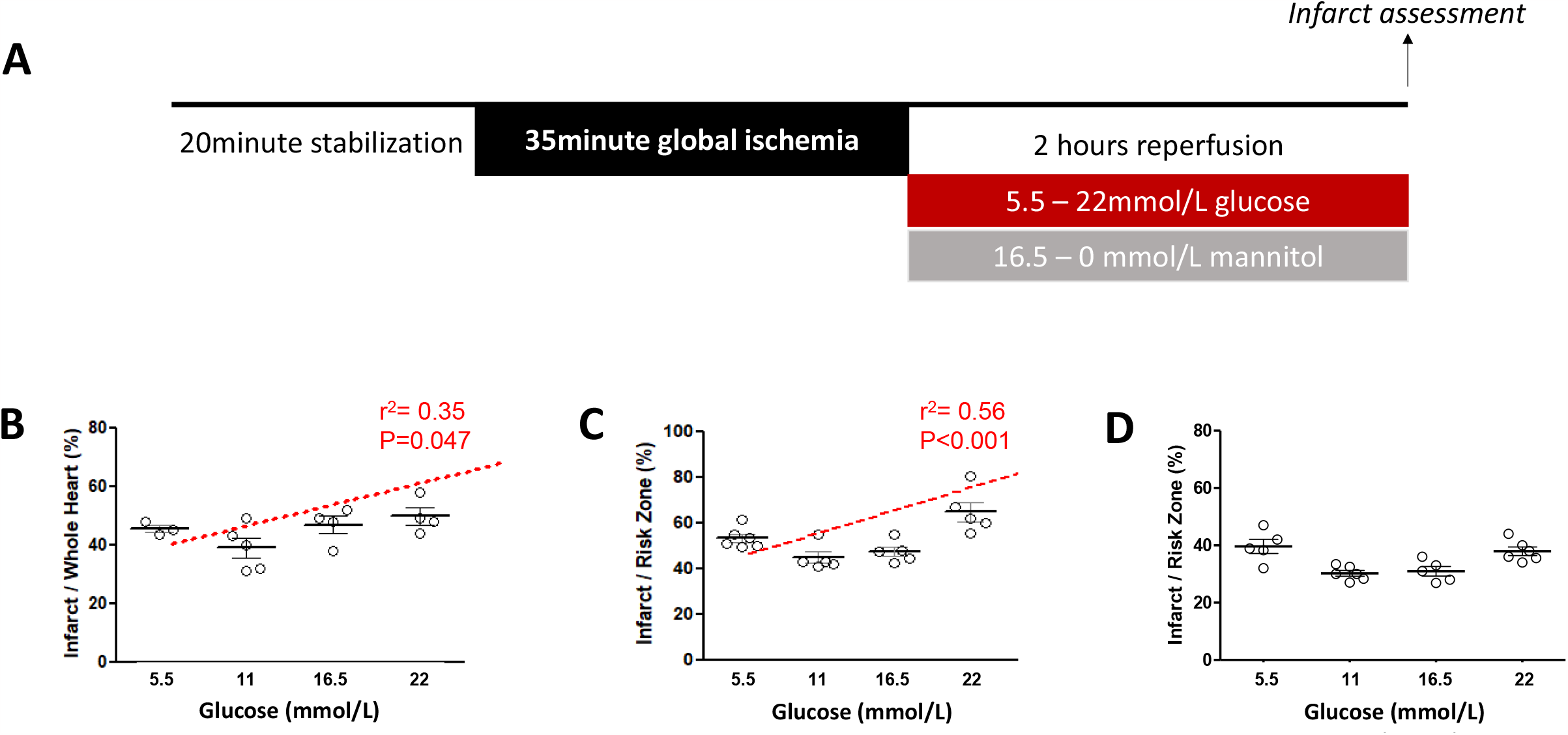
Glucose-infarct size dose-response in non-diabetic and diabetic mouse and rat heart models. **A:** Protocol for *ex-vivo* experiments. **B:** C57BL/6J non-diabetic mouse hearts (n=4/group) revealed a J-shaped dose-response curve, with minimum and maximum at 11mmol/L and 22mmol/L glucose respectively. A linear dose-response was observed with incremental glucose, with an r^2^=0.35, p=0.047. **C:** SD non-diabetic rat hearts (n=6/group) had minimum and maximum infarction at 11mmol/L and 22mmol/L glucose respectively, with a linear dose-response relationship between these values (r^2^=0.56, p=0.0009). **D:** GK diabetic rat heart (n=6/group) had a blunted dose-response curve compared to non-diabetic hearts, with a right-shift of the dose-response, with minimum infarction occurring at 16.5mmol/L and attenuated peak infarct sizes.

### Isolation of adult rat cardiomyocytes

Cardiomyocyte isolation was based on a previous protocol with minor modifications^16^. Rat hearts were sequentially perfused with cell isolation buffer (130mmol/L NaCl, 5.4mmol/L KCI, 1.4mmol/L MgCl_2_, 0.4mmol/L Na_2_HPO_4_, 4.2mmol/L HEPES, 10mmol/L glucose, 20mmol/L taurine, and 10mmol/L creatine, pH 7.4) containing i) 750 μmol/L CaCl_2_, ii) 50μg/ml EGTA, and iii) 100μmol/L CaCl2 + 50mg/dL Collagenase Type 5. Ventricles were minced and triturated with Collagenase 5 (50mg/dL) and Protease XIV (5mg/dL) with the resulting cells pelleted by gravity and resuspended in cell isolation buffer containing 1% BSA and increasing concentrations of CaCl2 up to a final concentration of 1mM.

### Tissue collection and processing

Heart, kidney, proximal gut, and skeletal muscle samples were excised from rats under terminal anaesthesia and washed in ice-cold Phosphate-buffered saline (PBS). For RT-PCR, tissue samples were stored in RNAlater™. For analysis of tissues by immunohistochemistry and RNAscope, samples were fixed in 4% paraformaldehyde overnight, washed with PBS and stored in 70% ethanol until processed for paraffin-embedding and sectioning.

### Polymerase chain reaction (PCR)

RNA was extracted from tissue and cells using RNeasy Mini Kit and concentration and purity measured by 260/280 OD ratio. RNA (1μg) was reverse-transcribed into cDNA using oligo dT Primers and an RT–PCR Kit. PCR was performed for *Sglt1* (forward primer 5’-TGCGGGGTCCACTATCT-3’; reverse primer 5’-CAACTCCAGAGTCGCCA-3’) and *Sglt2* (forward primer 5’-TTCTGTCATCGCACTCTTGG-3’; reverse primer 5’-GATCCTTGGACACCGTCAGT-3’).

### RNAscope

RNAscope on formalin-fixed paraffin-embedded (FFPE) sections (5μm) were performed using the Multiplex Fluorescent Assay kit as per manufacturer’s instructions. Sections were hybridized with probe-sets targeting *Sglt1* or *Sglt2* mRNA and counterstained with 4’,6-diamidino-2-phenylindole (DAPI). Positive and negative probes were used as controls. Co-detection of ***Sglt1*** and ***Sglt2*** mRNA with anti-CD31 antibodies (Abcam; ab222783,1:150) followed by Goat Anti-Rabbit IgG H&L (Alexa Fluor®488; ab150081,1:300) or the cell marker Wheat Germ Agglutinin (WGA), Alexa Fluor™ 488 Conjugate (Invitrogen™; W11261,1:150) was performed according to manufacturer’s instructions. For semiquantitative analysis of RNA signals, the number of DAPI-stained nuclei and *Sglt* mRNA signals were ascertained in five random microscopic fields of each section using Image J. Average RNA signals for each tissue section was determined by dividing the total section *Sglt* mRNA signal count by total section nuclei count.

#### Immunohistochemistry

Immunohistochemical probing of FFPE sections were carried out as previously described^17^ with a primary antibody against SGLT1 (ab14685, Abcam) and counterstained in Mayers hematoxylin.

#### Single-nucleus RNA-sequencing analysis of human hearts

To profile the expression of SGLT-encoding transcripts in the human heart, we analysed two publicly available single-nucleus RNA-sequencing (snRNA-seq) datasets sourced from the Gene Expression Omnibus (GSE183852)^18^ and the Broad Institute’s Single Cell Portal (https://singlecell.broadinstitute.org/single_cell/study/SCP498/transcriptional-and-cellular-diversity-of-the-human-heart)^19^. Analyses were performed in R (v4.0.2), using the Seurat package^20^. An object was created from each dataset, containing donors without evidence of overt cardiac disease, with associated metadata including biological sex or age group of the donors^18^ and cardiac chamber from which the cells originated from^19^. Cardiomyocyte count matrices were normalised and scaled before principal component analysis, with donor integration using Harmony^21^, shared nearest neighbor graphing, and unsupervised clustering. Transcript enrichment was visualised using Feature plots or Violin plots and quantified using differential expression tests. The code used to perform analysis can be accessed at: https://github.com/daniyal-jafree1995/collaborations/blob/main/Almalki%26Arjun_SGLT.R.

#### Statistical analysis

Differential expression tests for snRNA-seq data utilised Wilcoxon Rank Sum tests in RStudio. All other analyses were performed using GraphPad Prism version 6. Normality was assessed using the Shapiro–Wilk test. Normally distributed data is presented as mean ± SEM and analysed by unpaired t-test for 2 independent groups and ANOVA with Tukey’s multiple comparison test for 3 or more independent groups. Following preliminary experimental data, power calculations were performed *a priori*, to calculate group size in subsequent studies, based on achieving a power of 80% and an α of 0.05. *P* value of <0.05 was considered significant.

## Results

### Elevated glucose during reperfusion leads to increased myocardial infarct size

We hypothesised that elevated glucose at the time of reperfusion following injurious ischemia, would result in increased myocardial injury. To test this, we increased the glucose content of the perfusion buffer, with cross-group osmolality maintained with mannitol (**Fig.1A**). In both C57Bl6 mice and SD rats (**Fig.1B,C** respectively), the smallest infarct size was observed when the heart was perfused with 11mmol/L glucose and increased in a linear fashion with increasing glucose concentration, consistent with previous data^22^. At glucose concentrations below 11mmol/L, there was a trend towards an increased infarct size, commiserate with the “J”-curve observed in ACS patients. These preliminary dose-ranging studies were not powered to detect this increase as being significant. However, high glucose at the time of reperfusion following injurious ischemia is injurious in a dose-dependent fashion, increasing infarct size with an r^2^ of 0.35 and 0.56 (p<0.05 and p<0.001) in mouse and rat respectively (**Fig.1B,C**).

Similar to mortality outcomes seen in human patients, the relationship between glucose concentration and infarct size was significantly blunted in diabetic Goto-Kakizaki rat hearts (**Fig.1D**). There is a numerically, but not statistically larger infarct in low and very high glucose levels, but this experiment was not powered to determine significance in glucose-mediated cardiac injury in diabetic rat. Nonetheless, there would appear to be a right-shift of the “J-curve” dose-response curve seen in diabetic heart *versus* that seen in the non-diabetic heart: the smallest infarct in diabetic heart was observed at 16.5mmol/L rather than 11mmol/L in non-diabetic heart.

### Expression of SGLT transcripts in the myocardium: SGLT1 but no SGLT2

We assessed tissue expression of *SGLT1* and *SGLT2* mRNA, using kidney as a positive control and skeletal muscle as a negative control tissue. Heart clearly expressed SGLT1, but not SGLT2 (**Fig.2A** and **Fig.S1**). The myocardium SGLT1 expression was qualitatively less than that seen in kidney, consistent with published data^23^, with SGLT1 expression in ventricular and atrial chambers (**Fig.S2**).

**Figure 2:**
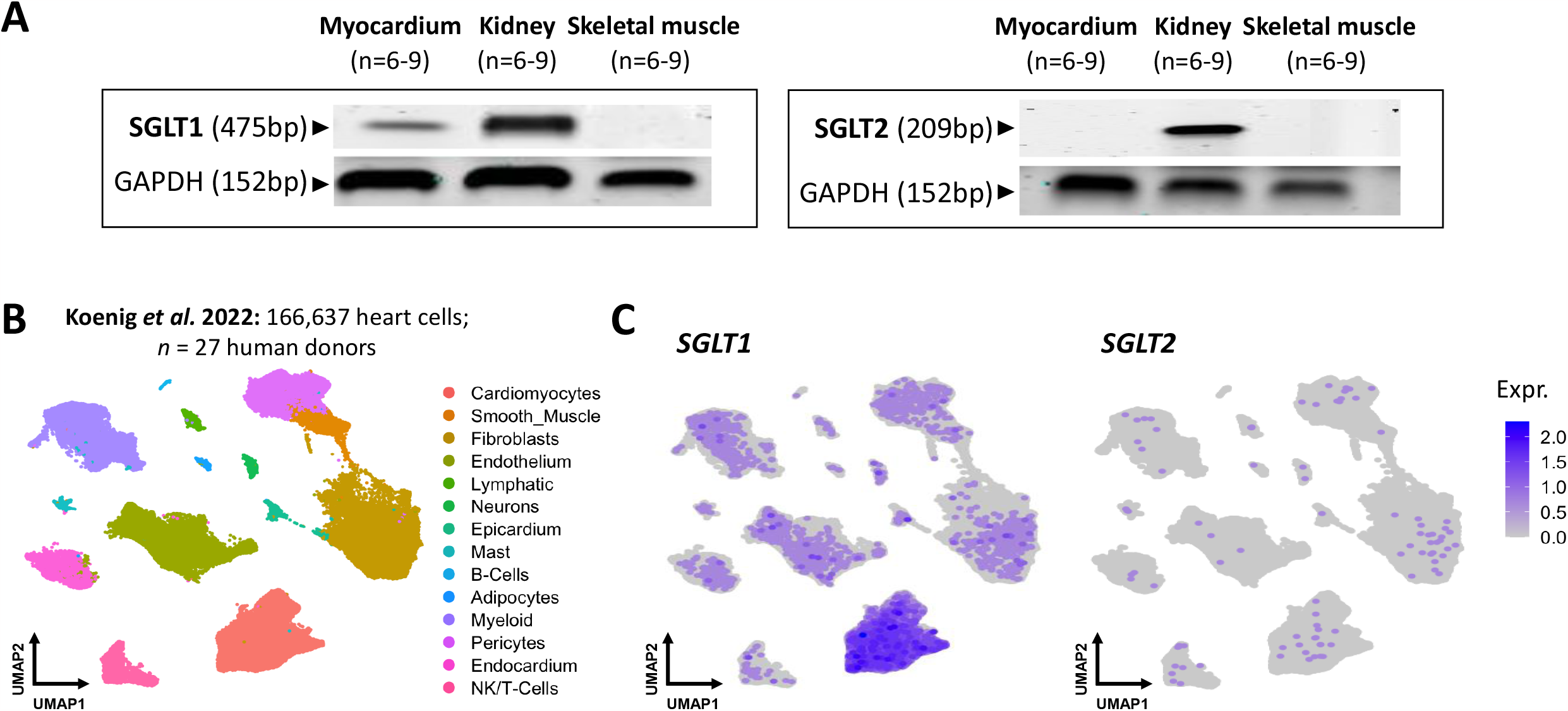
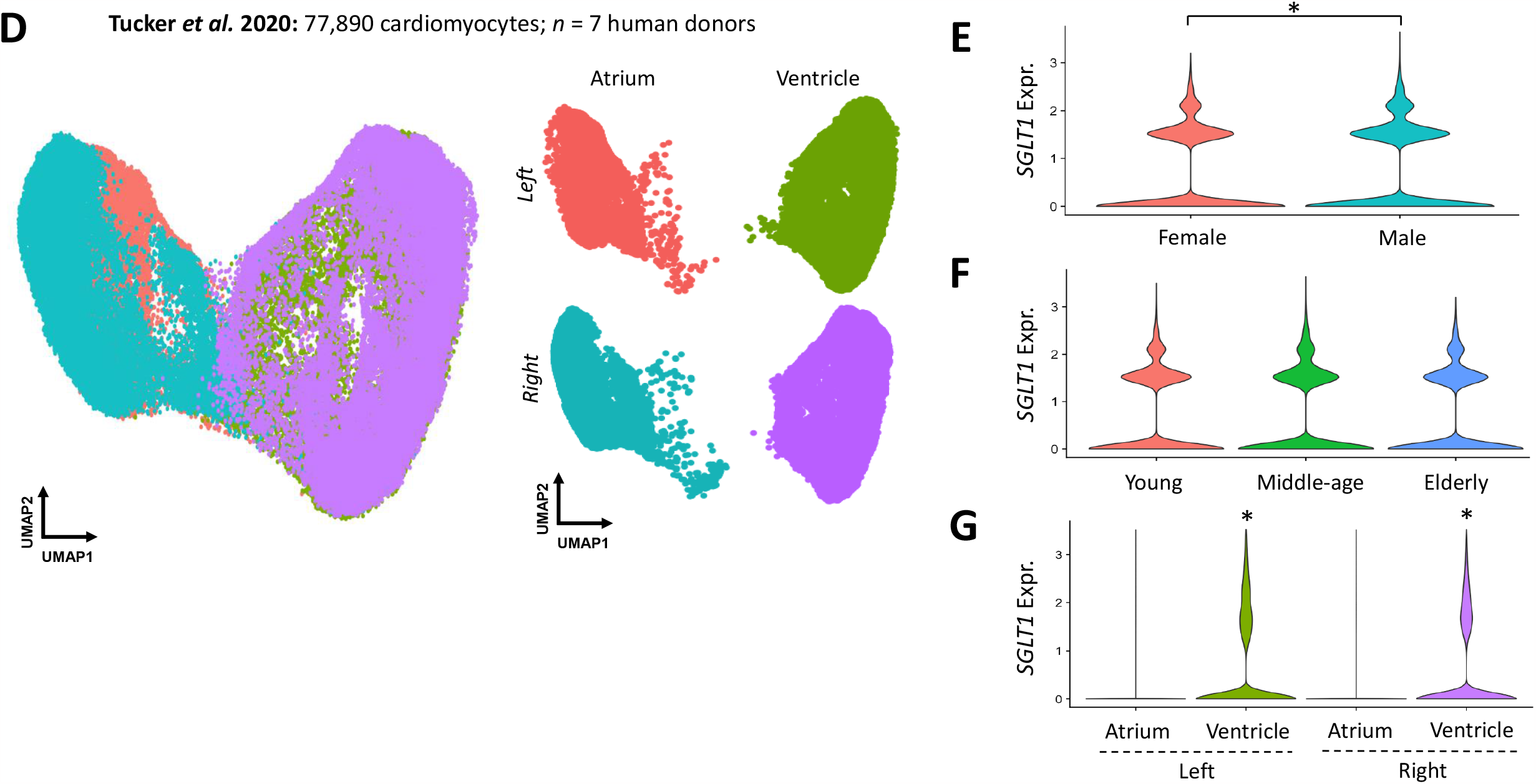
SGLT1 and SGLT2 expression in healthy rat and human myocardium. **A:** *SGLT1* and *SGLT2* rtPCR in rat heart, kidney (positive control) and skeletal muscle (negative control). In heart, *SGLT1* but not *SGLT2* mRNA is detected. **B:** Uniform manifold approximation and projection (UMAP) of snRNA-seq data of human heart cells from Koenig *et al*. 14 distinct cell types were identified, including cardiomyocytes. **C:** *SGLT1* expression (left UMAP) is enriched in cardiomyocytes. Conversely, *SGLT2* expression (right UMAP) is scant. **D:** UMAP of snRNA-seq data human cardiomyocytes (top UMAP) from Tucker *et al*. The bottom UMAP represents stratification of cardiomyocytes into the four chambers, with atrial and ventricular cardiomyocytes resolving separately. Analysis of the relative expression of *SGLT1* between female and male (**E**), in different age-groups (**F**) and in atrial *versus* ventricular cardiomyocytes (**G**).

We then determined the cellular expression profiles of SGLT-encoding transcripts within the human heart, leveraging two snRNA-seq datasets. One dataset comprised of 166,637 heart cells featuring fourteen cell types from 27 donors^18^ (**Fig.2B**) revealed myocardial enrichment for *SGLT1* (average log fold change; log_2_FC = 0.64, adjusted *p*<0.0001) with basal expression in non-myocardial cell types (**Fig.2C**). *SGLT2* expression was scant in the human heart (**Fig.2C**). To validate this in an independent dataset, we interrogated snRNA-seq data of 77,890 atrial or ventricular cardiomyocytes from seven human donors^19^ with *SGLT1* but not *SGLT2* expressed, the former enriched within cardiomyocytes (log_2_FC = 1.02, adjusted *p*<0.0001). Donors within the datasets were stratified by biological sex, age or heart chamber of origin (**Fig.2D**). Our analyses demonstrated minor variability of myocardial *SGLT1* expression across individuals at different age groups, albeit with subtle but statistically significant enrichment in male donors (log_2_FC = 0.13, adjusted *p*=1.3 × 10^-16, **Fig.2E,F**). Moreover, *SGLT1* was enriched in ventricular compared to atrial cardiomyocytes (**Fig.2G**).

### RNAscope and immunohistochemistry

RNAscope was used to visualise SGLT1 expression in the rodent heart and kidney (probe negative and positive controls together with small intestine and skeletal muscle positive and negative tissue controls shown in **Fig.S3-5**). The whole myocardium had clearly demonstrable *SGLT1* expression within the vasculature and cardiomyocytes (**Fig.3A**). No myocardial expression was observed with *SGLT2*, although there is clear expression in the kidney (**Fig.3B**). The vascular expression of *SGLT1* appears to be predominantly endothelial, as determined with counterstaining with WGA and CD31 (**Fig.3C,D**). Immunohistochemistry with *SGLT1* antibody was consistent with the mRNA expression data, with staining within vascular structures and cardiomyocytes (**Fig.3E**).

**Figure 3:**
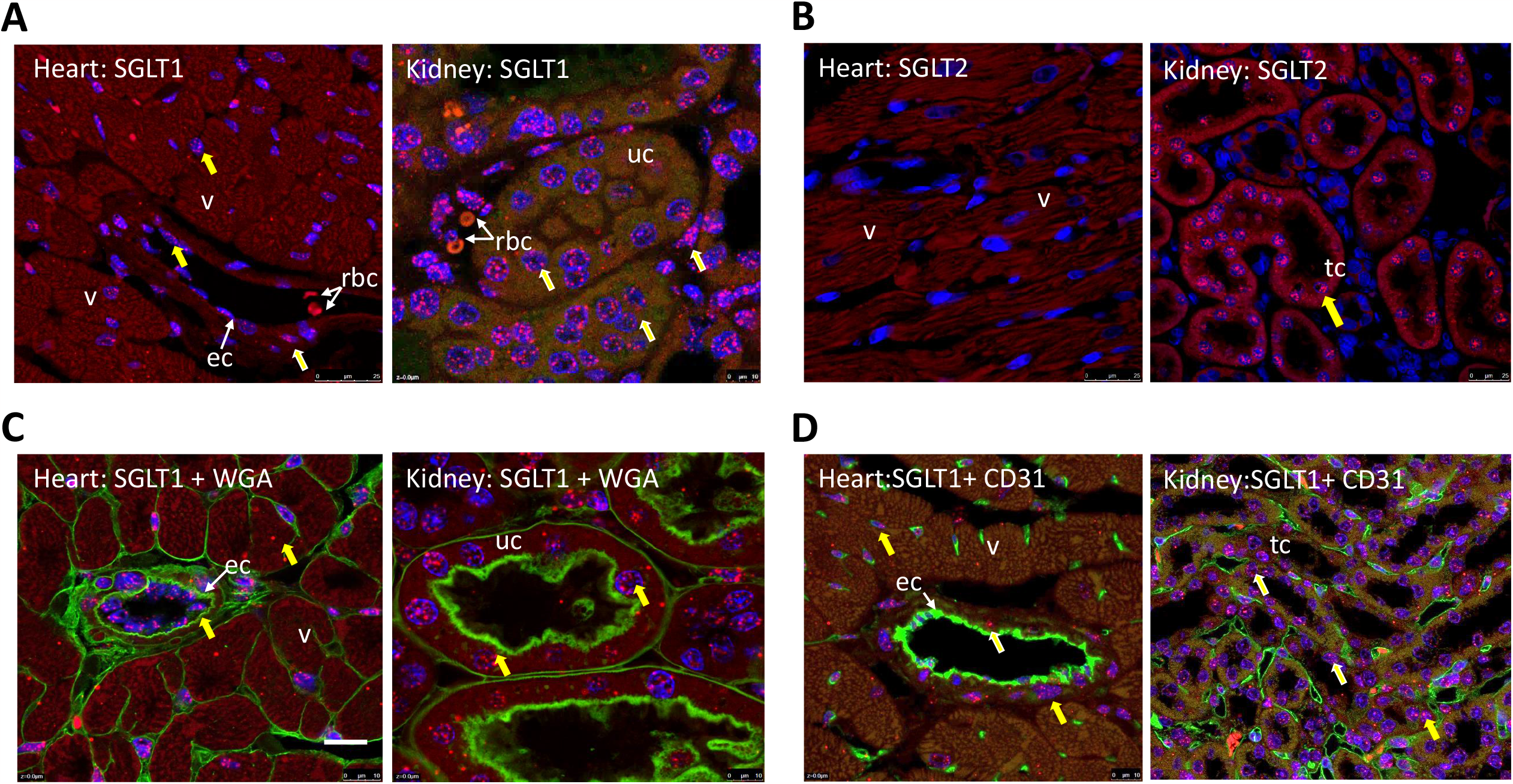
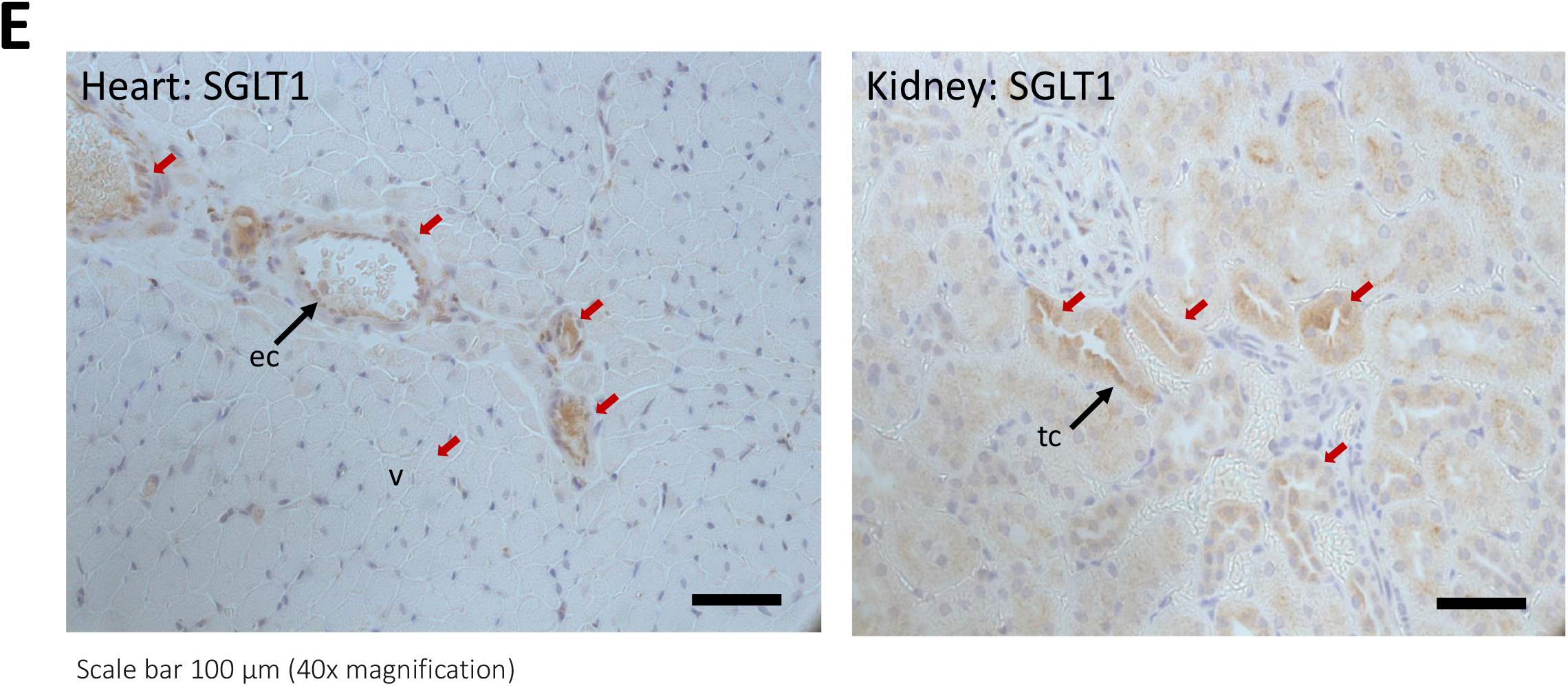
SGLT1 and SGLT2 expression in the myocardium. **A:** RNAscope revealed *SGLT1* mRNA expression in both cardiomyocytes (which auto-fluoresce red), and vascular structures. Kidney is a positive control, with a strong SGLT1 signal within the proximal nephron. *(v) = ventricular cardiomyocyte; (ec) = endothelial cell; (rbc) = red blood cell; (tc) = tubular cell. Yellow arrows highlight SGLT detection signal*. **B:** There was no SGLT2 signal detectable within the heart, in contrast to kidney. Vascular markers, cell membrane marker, WGA (**C**), and endothelial marker CD31 (**D**) were used to show SGLT1 distribution. **E:** Immunohistochemistry for SGLT1 protein expression reveals similar cellular staining pattern (brown, highlighted with red arrows) in heart and kidney to that seen with RNAscope. In the heart, staining can be found in the sarcolemma of the cardiomyocytes and the endothelium of arterioles and capillary structures.

### Impact of non-selective SGLT inhibition upon glucose associated excess injury

We then examined whether SGLT isoforms mediate the augmented myocardial injury associated with raised reperfusion glucose using three pharmacologically distinct inhibitors: specific, non-selective SGLT inhibitor phlorizin, SGLT2 inhibitor, canagliflozin and SGLT1inhibitor, mizagliflozin (**Fig.4A**).

**Figure 4:**
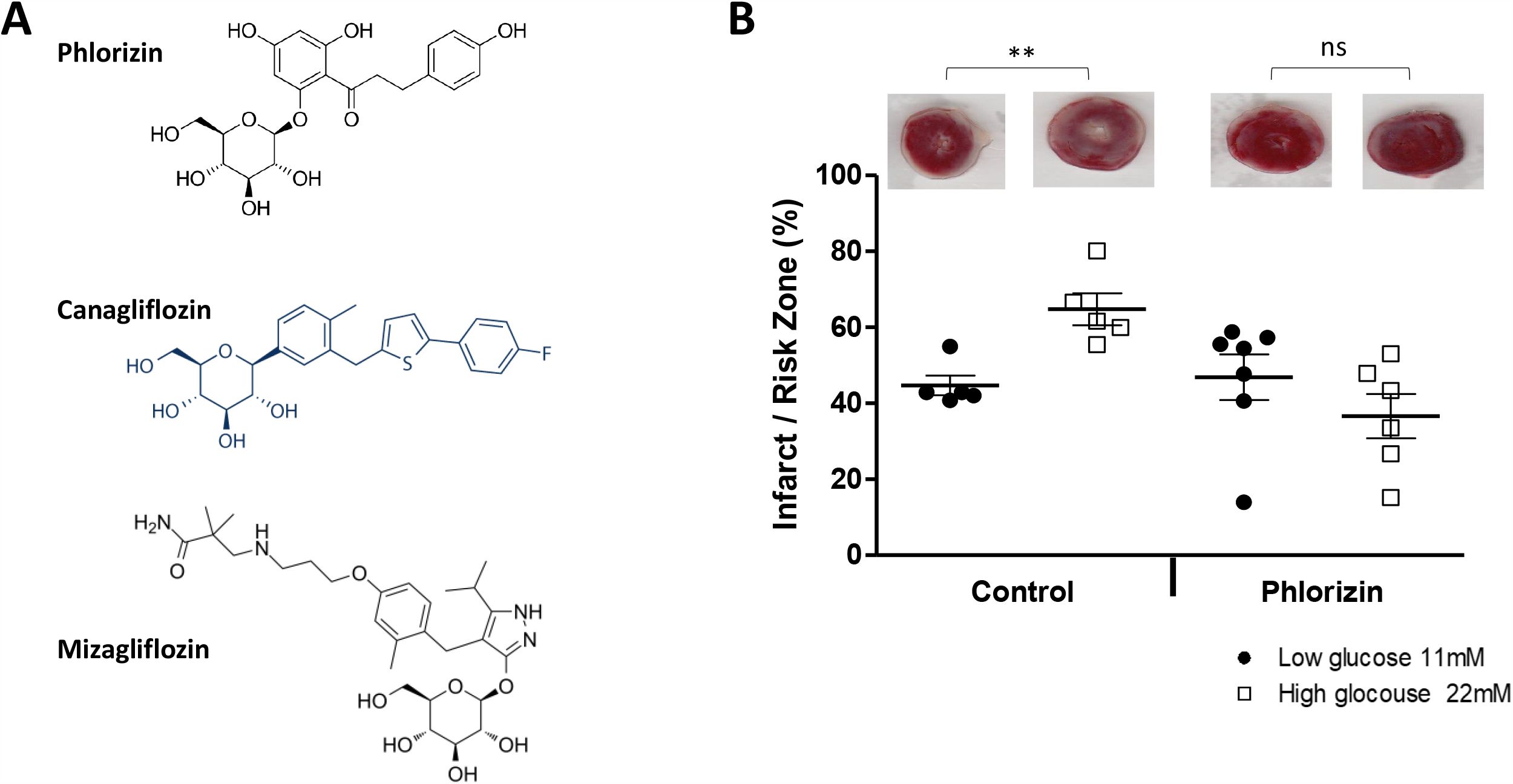
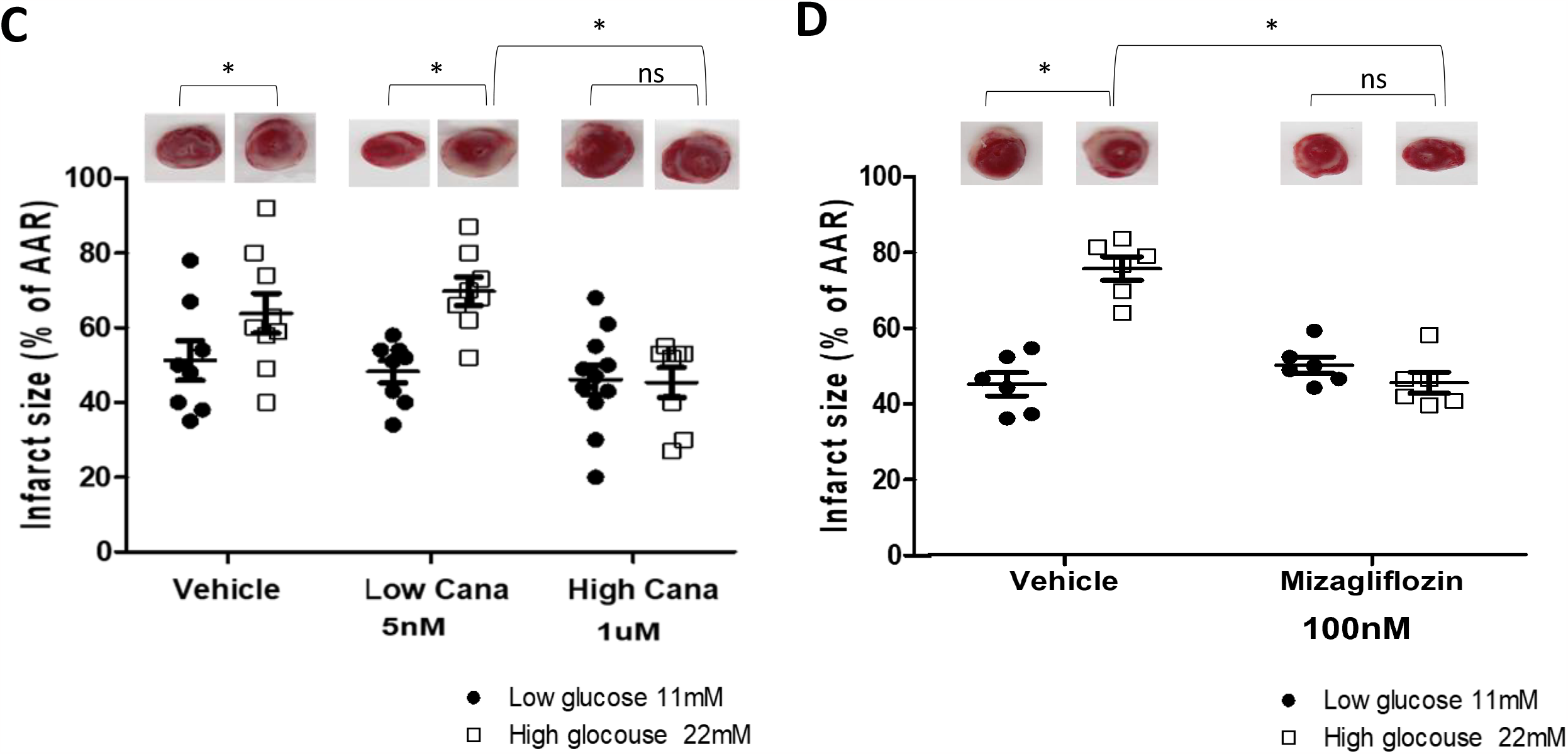
Inhibition of SGLT2 and SGLT1 in 11 and 22mmol/L glucose at reperfusion. **A:** Chemical structures of the SGLT inhibitors used. **B:** In non-diabetic heart, elevated glucose at reperfusion results in a significant increase in infarct over size the control condition which is completely abrogated by the administration of phlorizin (3 μmol/L). ** p<0.001 22mmol/L versus 11mmol/L reperfusion glucose. **C:** There is a significant increase in infarct size with high (22mmol/L) glucose versus 11mmol/L, which is not ameliorated by 5nmol/L canagliflozin (Cana). However, 1μmol/L canagliflozin abrogates this excess injury. **D:** Mizagliflozin, abrogates the excess injury associated with elevated glucose. Representative mid-ventricular myocardial slices shown. * p<0.05 versus respective control.

Phlorizin (3μmol/L) was added with the elevated (22mmol/L) glucose or standard glucose (11mmol/L + 11mmol/L mannitol) at the time of reperfusion. The EC_50_ of phlorizin in rat for SGLT2 and SGLT1 is 75 and 302nmol/L respectively^24^, and GLUT transport is only 10% inhibited at 20μmol/L^24^, thus 3μmol/L would be expected to inhibit both SGLT2 and SGLT1 without significant impact upon GLUT transport. As previously found, high glucose, led to a significant 44% increase in infarct size in DMSO control heart (from 45±2.6% to 65±4.2%, p<0.01), which was completely abrogated by the administration of phlorizin (65±4.2% to 37±5.8% in 22mmol/L with and without phlorizin respectively, p<0.01, **Fig.4B**). Phlorizin had no impact upon infarct size under “standard” glucose conditions of 11mmol/L (45±2.6% *versus* 47±5.8% in control versus phlorizin). Therefore, phlorizin is only cardioprotective under conditions of elevated glucose.

### Selective SGLT1 versus SGLT2 inhibition

Canagliflozin is marketed as an SGLT2 inhibitor, but compared to other SGLT2 inhibitors, the relative potency for SGLT2 over SGLT1 inhibition is less, with IC_50_ of 4.2nmol/L and 663nmol/L respectively in man, and 3.7nmol/L and 555nmol/L in rat. The *in-vitro* IC_50_ for canagliflozin against GLUT isoform activity is >1000nmol/L^25^. Thus, using 5nmol/L will achieve selective SGLT2 inhibition and 1μmol/L combined SGLT2 + SGLT1 inhibition, and neither will inhibit GLUT glucose transport. We hypothesised that SGLT2-specific dose canagliflozin would not impact infarct size, whereas 1μmol/L, inhibiting both SGLT1 and SGLT2, would abrogate the excess injury associated with high glucose during reperfusion. As part of our statistical design, we took the data from our phlorizin study to make a calculation of group size. To achieve an 80% power and an α of 0.05, we calculated a sample size of n=9 per group to determine a difference of 25%.

In vehicle controls, high glucose increased infarct size (from 51±5.3% to 64±5.3%, p<0.05, **Fig.4C**). 5nmol/L canagliflozin failed to ameliorate the increase in infarction associated with high glucose (48±2.9% versus 70±3.8% in 11 and 22mmol/L glucose respectively, p<0.001), however 1μmol/L canagliflozin completely abrogated the excess injury associated with 22mmol/L glucose (45±4.0% versus 46±4.0% for 11 and 22mmol/L glucose respectively, p=0.99, **Fig.4C**, representative heart slices **Fig.S6**). Thus, specific SGLT2 inhibition had no impact upon the excess injury associated with high glucose, whereas combined SGLT2+SGLT1 inhibition abrogated this deleterious effect.

In the absence of demonstrable SGLT2 expression within the heart, the high 1μmol/L canagliflozin can be expected to be mediating its protective effect against high glucose mediated injury via myocardial SGLT1 inhibition. However, to further demonstrate the importance of SGLT1, we tested a specific SGLT1 inhibitor, Mizagliflozin which has an IC_50_ of 166nmol/L in rat^26^ and a Ki of 27.0nmol/L and 8170nmol/L for SGLT1 and SGLT2 respectively from *in-vitro* human studies^27^. 100nmol/L mizagliflozin will inhibit SGLT1 specifically, and in doing so, we hypothesised that it would completely abrogate glucose-mediated excess injury.

22mmol/L glucose increased infarct size relative to 11mmol/L (65±2.6% compared to 39±2.4%, p<0.001), but this excess injury was abrogated by mizagliflozin (43±1.9% versus 39±2.4% in 11 and 22mmol/L glucose respectively, p=0.65, **Fig.4D**, representative heart slices **Fig.S7**).

### Impact of diabetes upon SGLT1 and SGLT2 expression in the myocardium

Injury resulting from high glucose at the time of reperfusion, as found in human patient data, is attenuated in diabetic heart (**Fig.1D**). SGLT2 kidney expression increases with diabetes and published data indicates that diabetes alters myocardial SGLT1 expression. However, the impact of diabetes upon myocardial SGLT2 expression is unknown.

Myocardial SGLT expression was investigated by RNA-scope using ZDF and ZL rat hearts with the same animals’ kidney as a positive control tissue. In ZDF rat hearts, SGLT1 expression was 50% less than in non-diabetic heart (p<0.05, **Fig.5A**). In contrast, renal SGLT1 expression in ZDF and ZL rats were similar (**Fig.5B**). Conversely, SGLT2 expression remained absent in diabetic heart (**Fig.5C**) but was significantly increased by 50% in diabetic kidney relative to non-diabetic animals, as previously reported^28^ (**Fig.5D**).

**Figure 5:**
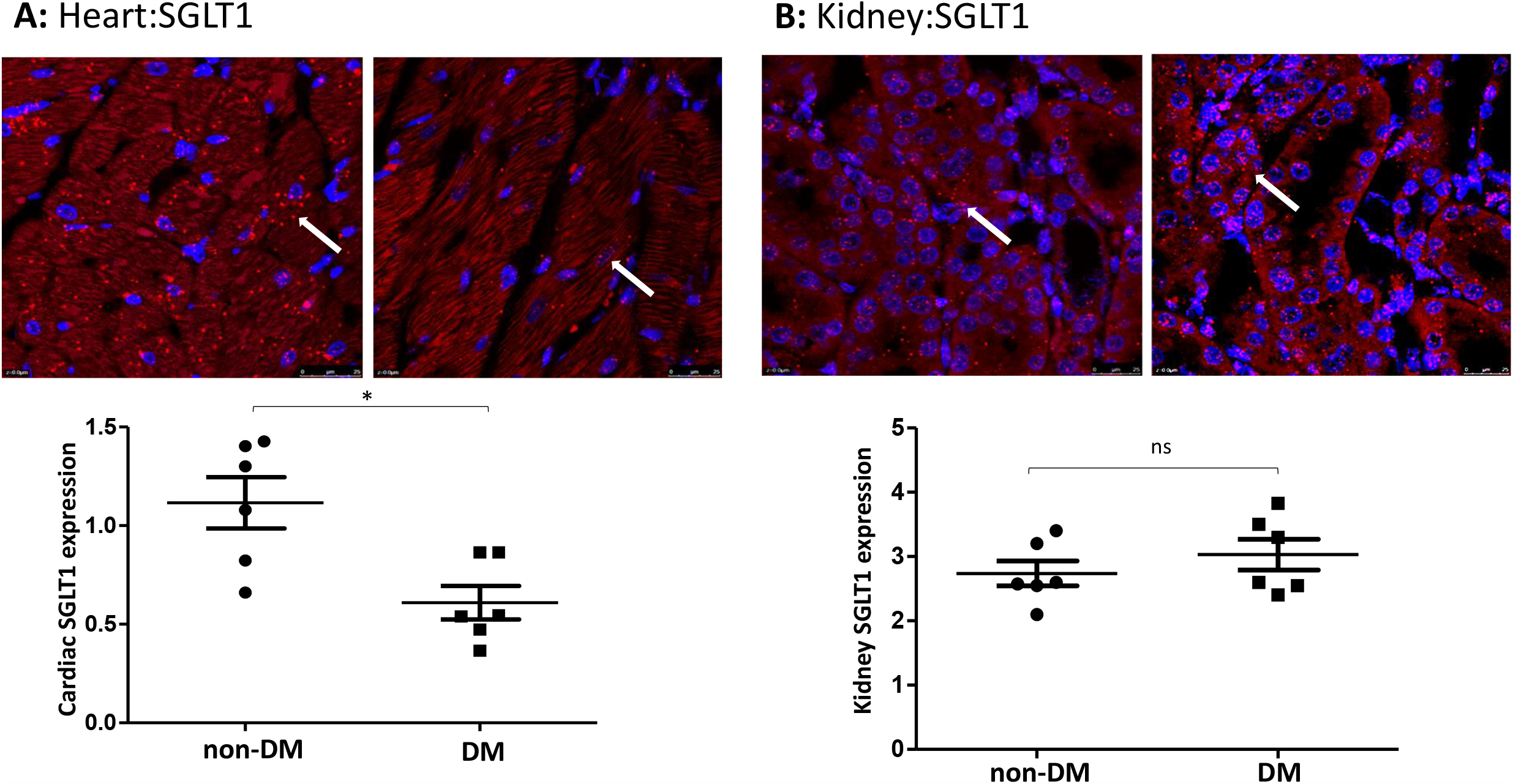
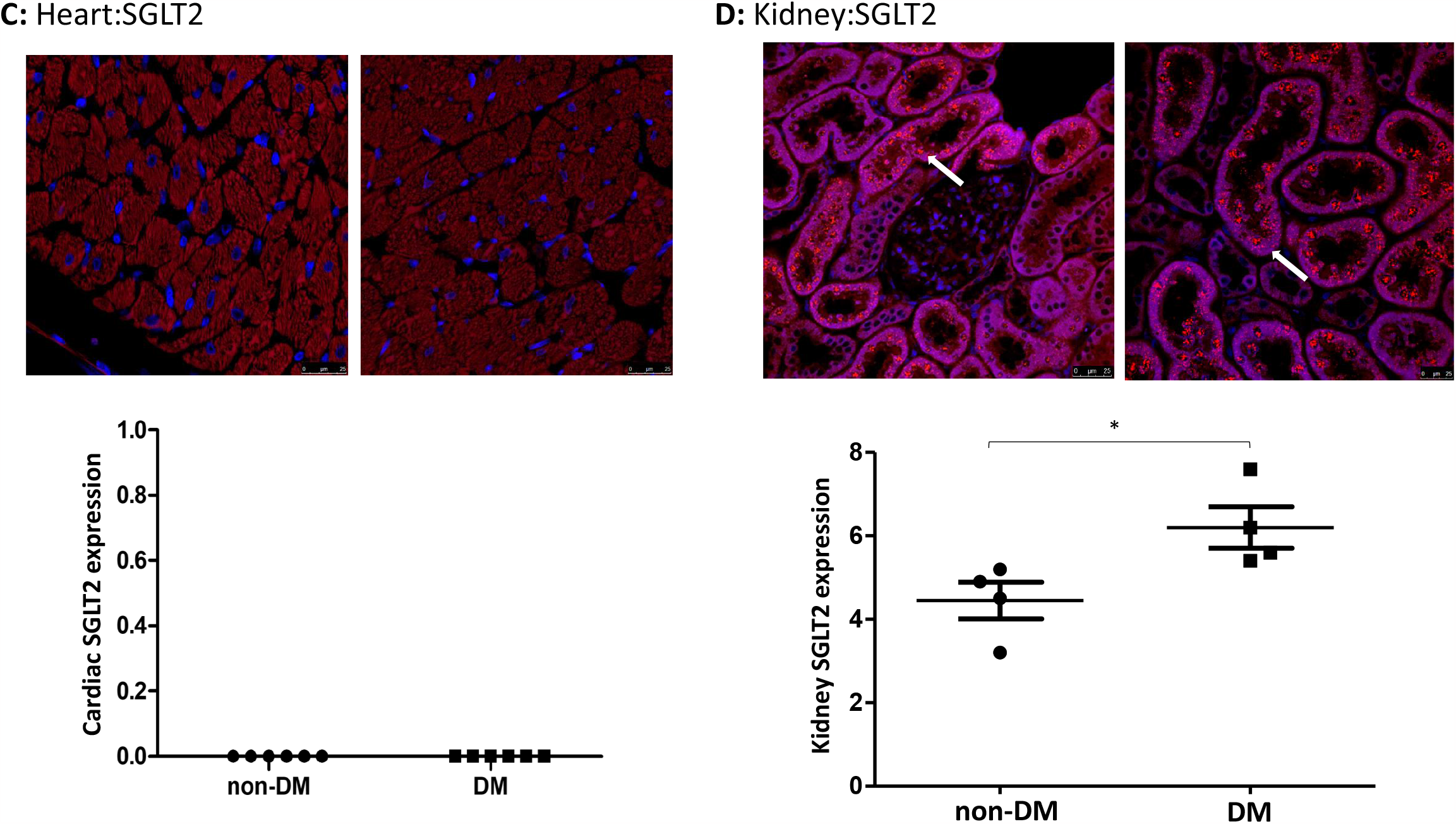
SGLT1 and SGLT2 expression in non-diabetic and diabetic heart. **A:** In ZL rat heart, myocardial *SGLT1* mRNA expression was markedly higher than that seen in ZDF rat heart (n=6/group). **B:** *SGLT1* mRNA expression in kidney was not significantly altered in the ZDF diabetic model. **C:** Diabetes did not result in a compensatory increase of *SGLT2* mRNA expression. **D:** Renal SGLT2 expression is significantly increased in diabetic kidney versus non-diabetic kidney. *p<0.05.

## Discussion

In this study, we demonstrate, for the first time, that elevated glucose with reperfusion, results in a dose-dependent increase in myocardial infarct size in mouse and rat hearts, thus demonstrating a “glucose paradox”. As with calcium, oxygen and pH upon reperfusion, rapid restoration of energy substrate availability following injurious ischemia should be beneficial, but a higher-than-normal glucose level at the time of reperfusion drives further injury, and we demonstrate that this is driven primarily through myocardial SGLT1 activity.

There is a striking similarity between our data, to that observed in ACS patients. In our model, as in patient mortality data, there is a J-shaped dose-relationship between glucose concentration and outcome, where excess infarction in the animal models and mortality in clinical studies is observed in both low and elevated glucose states. This similarity may be coincidental but given that larger infarcts are associated with poorer cardiovascular outcomes following AMI, there is a clear and testable hypothesis that can be derived: effectively managing glucose, particularly in patients without prior known diabetes, should result in better cardiovascular outcomes. Furthermore, our observation that heart of diabetic rat is resistant to high glucose also mirrors patient epidemiological data – somehow, the diabetic heart is relatively protected against the injurious impact of high glucose following AMI.

We find that SGLT1 is widely expressed within the rodent heart with high levels within the coronary arteriolar endothelium and cardiomyocytes. This alludes to an unexplored role of SGLT1 in the physiological transport of glucose in the myocardium, likely complimenting GLUT transporters in coronary blood glucose extraction and transport to the highly metabolically active myocytes. In this study, we have examined the pathophysiological setting of acute injurious myocardial ischemia and reperfusion. Inhibiting SGLT1 abrogates the excess myocardial injury associated with reperfusion elevated glucose, but under standard “normal” 11mmol/L glucose conditions, SGLT1 inhibition has no impact upon infarct size. These data are important in terms of translatability. Hyperglycemia is common in ACS and is poorly managed owing to concerns of inducing hypoglycemia that can be as deleterious to mortality outcome as hyperglycemia. SGLT inhibitors do not result in hypoglycemia, and thus offer the potential for safely normalising glucose. Moreover, we show that the protective effect of SGLT1 inhibition is not reliant upon glucose normalisation: SGLT1 inhibition protects the heart despite persisting high glucose. Therefore, inhibiting SGLT1 has two beneficial modes of action: a direct anti-glucotoxic and a secondary glucose-lowering effect. The diabetic heart is resistant to high glucose, and we find the glucose/injury dose-response curve appears both blunted and right-shifted (**Fig.1D**). SGLT1 appears pivotal to glucose-mediated injury in non-diabetic heart, it is therefore notable that in our model, the diabetic heart has significantly lower SGLT1 expression. This suggests that in diabetes, SGLT1 expression is regulated to some degree by circulating glucose levels; it seems plausible to hypothesise that the diabetic heart, with higher circulating glucose, down-regulates SGLT1 in response to energy “plentifulness”. Under the pathophysiological conditions of AMI, SGLT1 downregulation protects the diabetic heart against injurious impact of elevated glucose.

This regulation of SGLT1 has important implications. When designing clinical studies to manage hyperglycemia in ACS, one has to be wary of the diabetic status. A large diabetic population will both reduce the expected injury and attenuate the benefit of any glucose-moderating intervention, and the trial would need to be powered accordingly. Our data also suggests that glucose-mediated injury occurs rapidly (within two hours of reperfusion) – so delays in achieving effective glucose lowering, if not targeting SGLT1 directly, would likely lead to failure to protect the heart – and this may explain why intensive insulin therapy studies such as DIGAMI-2 and HI-5 failed to achieve expectations.

Others have suggested that SGLT1 expression is altered by disease, including in the setting of diabetes. However, there is some controversy in this area: Banerjee et al demonstrated reduced SGLT1 expression in a streptozotocin-induced type 1 diabetic mouse model, whereas they found increased expression in the *ob/ob* obese mouse model of type 2 diabetes. The explanation for the latter is unclear: the *ob/ob* mouse is similar to our ZDF rat, with both models having leptin signalling deficiency. However, heart failure (from a diabetic cardiomyopathy for example) or elevated leptin levels are both associated with increased SGLT1 expression^14^ – and these observations may in themselves have significant clinical importance, as hearts expressing more SGLT1 would be predicted to be more vulnerable to the adverse impact of elevated glucose and thus would likely benefit even more from SGLT1 inhibition, if presenting with AMI.

### Study limitations

Our study has a reductionist experimental design using the *ex-vivo* Langendorff heart perfusion model giving us full control over glucose concentration, the timing of raised glucose and duration of ischemia and onset of reperfusion. With crystalloid perfusates, such as the modified Krebs-Henesleit buffer used here, there are no concerns regarding the viscosity and thus flow of the perfusate through the coronary vasculature. However, plasma viscosity is a likely contributing factor in microvascular obstruction and no-reflow, would be a concern in the whole animal and in patients. Stress hyperglycemia remains relatively understudied, and therefore adequately and reproducibly replicating these conditions is difficult. A possible avenue is intravenous administration of glucose, but this may have compensatory impacts upon circulating insulin and gut hormone release, that may have unpredictable effects upon myocardial injury and SGLT1 expression. However, given that the reductionist model we have used have led to such a remarkable dose-response similarity to the epidemiological data, assuming that infarct size correlates directly to mortality^29^, suggests that the model assumptions we have made are appropriate, and presents a platform for further studies.

The lack of reliable, validated commercial antibodies has meant we have been unable to quantify SGLT protein expression by Western blot analysis. However, our data on SGLT mRNA expression and distribution, and *ex-vivo* IR experiments confirming the functional impact of three, chemically distinct SGLT inhibitors, reveal both transcriptional and functional protein evidence of SGLT1 expression within the myocardium.

## Conclusion

We demonstrate for the first time that elevated glucose, at the time of reperfusion following an injurious ischemic insult, results in a biologically important 45% increase in myocardial injury and infarction. This relationship is marked in the non-diabetic heart but is significantly attenuated in the diabetic heart, which appears correlated with suppressed diabetic myocardial SGLT1 expression. We show that SGLT1 widely expressed throughout the myocardium, expressed in vascular endothelium as well as cardiomyocytes, and is critically linked to the development of reperfusion glucotoxicity. Our data therefore suggest an exciting opportunity to repurpose existing SGLT1 inhibitors to further improve outcomes in ACS with concomitant hyperglycemia.

## Data Availability

All data is available.

## Acknowledgements

- See funding sources below.

## References

1. Barquera S, Pedroza-Tobias A, Medina C, Hernandez-Barrera L, Bibbins-Domingo K, Lozano R, Moran AE. Global Overview of the Epidemiology of Atherosclerotic Cardiovascular Disease. Arch Med Res. 2015;46:328–338. doi: 10.1016/j.arcmed.2015.06.006

2. Deedwania P, Kosiborod M, Barrett E, Ceriello A, Isley W, Mazzone T, Raskin P. Hyperglycemia and acute coronary syndrome: a scientific statement from the American Heart Association Diabetes Committee of the Council on Nutrition, Physical Activity, and Metabolism. Circulation. 2008;117:1610–1619. doi: 10.1161/CIRCULATIONAHA.107.188629

3. Ritsinger V, Hagstrom E, Lagerqvist B, Norhammar A. Admission Glucose Levels and Associated Risk for Heart Failure After Myocardial Infarction in Patients Without Diabetes. J Am Heart Assoc. 2021;10:e022667. doi: 10.1161/JAHA.121.022667

4. Malmberg K, Ryden L, Hamsten A, Herlitz J, Waldenstrom A, Wedel H. Effects of insulin treatment on cause-specific one-year mortality and morbidity in diabetic patients with acute myocardial infarction. DIGAMI Study Group. Diabetes Insulin-Glucose in Acute Myocardial Infarction. Eur Heart J. 1996;17:1337–1344.

5. Malmberg K, Ryden L, Wedel H, Birkeland K, Bootsma A, Dickstein K, Efendic S, Fisher M, Hamsten A, Herlitz J, et al. Intense metabolic control by means of insulin in patients with diabetes mellitus and acute myocardial infarction (DIGAMI 2): effects on mortality and morbidity. Eur Heart J. 2005;26:650–661. doi: 10.1093/eurheartj/ehi199

6. Cheung NW, Wong VW, McLean M. The Hyperglycemia: Intensive Insulin Infusion in Infarction (HI-5) study: a randomized controlled trial of insulin infusion therapy for myocardial infarction. Diabetes Care. 2006;29:765–770. doi: 10.2337/diacare.29.04.06.dc05-1894

7. O’Gara PT, Kushner FG, Ascheim DD, Casey DE, Jr., Chung MK, de Lemos JA, Ettinger SM, Fang JC, Fesmire FM, Franklin BA, et al. 2013 ACCF/AHA guideline for the management of ST-elevation myocardial infarction: a report of the American College of Cardiology Foundation/American Heart Association Task Force on Practice Guidelines. Circulation. 2013;127:e362–425. doi: 10.1161/CIR.0b013e3182742cf6

8. Collet JP, Thiele H, Barbato E, Barthelemy O, Bauersachs J, Bhatt DL, Dendale P, Dorobantu M, Edvardsen T, Folliguet T, et al. 2020 ESC Guidelines for the management of acute coronary syndromes in patients presenting without persistent ST-segment elevation. Eur Heart J. 2021;42:1289–1367. doi: 10.1093/eurheartj/ehaa575

9. Zinman B, Wanner C, Lachin JM, Fitchett D, Bluhmki E, Hantel S, Mattheus M, Devins T, Johansen OE, Woerle HJ, et al. Empagliflozin, Cardiovascular Outcomes, and Mortality in Type 2 Diabetes. N Engl J Med. 2015;373:2117–2128. doi: 10.1056/NEJMoa1504720

10. Di Franco A, Cantini G, Tani A, Coppini R, Zecchi-Orlandini S, Raimondi L, Luconi M, Mannucci E. Sodium-dependent glucose transporters (SGLT) in human ischemic heart: A new potential pharmacological target. Int J Cardiol. 2017;243:86–90. doi: 10.1016/j.ijcard.2017.05.032

11. Sabolic I, Vrhovac I, Eror DB, Gerasimova M, Rose M, Breljak D, Ljubojevic M, Brzica H, Sebastiani A, Thal SC, et al. Expression of Na+-D-glucose cotransporter SGLT2 in rodents is kidney-specific and exhibits sex and species differences. Am J Physiol Cell Physiol. 2012;302:C1174–1188. doi: 10.1152/ajpcell.00450.2011

12. Zhou L, Cryan EV, D’Andrea MR, Belkowski S, Conway BR, Demarest KT. Human cardiomyocytes express high level of Na+/glucose cotransporter 1 (SGLT1). J Cell Biochem. 2003;90:339–346. doi: 10.1002/jcb.10631

13. Sawa Y, Saito M, Ishida N, Ibi M, Matsushita N, Morino Y, Taira E, Hirose M. Pretreatment with KGA-2727, a selective SGLT1 inhibitor, is protective against myocardial infarction-induced ventricular remodeling and heart failure in mice. J Pharmacol Sci. 2020;142:16–25. doi: 10.1016/j.jphs.2019.11.001

14. Banerjee SK, McGaffin KR, Pastor-Soler NM, Ahmad F. SGLT1 is a novel cardiac glucose transporter that is perturbed in disease states. Cardiovasc Res. 2009;84:111–118. doi: 10.1093/cvr/cvp190

15. Rossello X, Riquelme JA, He Z, Taferner S, Vanhaesebroeck B, Davidson SM, Yellon DM. The role of PI3Kalpha isoform in cardioprotection. Basic Res Cardiol. 2017;112:66. doi: 10.1007/s00395-017-0657-7

16. Shah M, He Z, Rauf A, Beikoghli Kalkhoran S, Heiestad CM, Stenslokken KO, Parish CR, Soehnlein O, Arjun S, Davidson SM, et al. Extracellular histones are a target in myocardial ischaemia-reperfusion injury. Cardiovasc Res. 2022;118:1115–1125. doi: 10.1093/cvr/cvab139

17. Long DA, Woolf AS, Suda T, Yuan HT. Increased renal angiopoietin-1 expression in folic acid-induced nephrotoxicity in mice. J Am Soc Nephrol. 2001;12:2721–2731. doi: 10.1681/ASN.V12122721

18. Koenig AL, Shchukina I, Amrute J, Andhey PS, Zaitsev K, Lai L, Bajpai G, Bredemeyer A, Smith G, Jones C, et al. Single-cell transcriptomics reveals cell-type-specific diversification in human heart failure. Nat Cardiovasc Res. 2022;1:263–280. doi: 10.1038/s44161-022-00028-6

19. Tucker NR, Chaffin M, Fleming SJ, Hall AW, Parsons VA, Bedi KC, Jr., Akkad AD, Herndon CN, Arduini A, Papangeli I, et al. Transcriptional and Cellular Diversity of the Human Heart. Circulation. 2020;142:466–482. doi: 10.1161/CIRCULATIONAHA.119.045401

20. Hao Y, Hao S, Andersen-Nissen E, Mauck WM, 3rd, Zheng S, Butler A, Lee MJ, Wilk AJ, Darby C, Zager M, et al. Integrated analysis of multimodal single-cell data. Cell. 2021;184:3573–3587 e3529. doi: 10.1016/j.cell.2021.04.048

21. Korsunsky I, Millard N, Fan J, Slowikowski K, Zhang F, Wei K, Baglaenko Y, Brenner M, Loh PR, Raychaudhuri S. Fast, sensitive and accurate integration of single-cell data with Harmony. Nat Methods. 2019;16:1289–1296. doi: 10.1038/s41592-019-0619-0

22. Owen P, du Toit EF, Opie LH. The optimal glucose concentration for intermittent cardioplegia in isolated rat heart when added to St. Thomas’ Hospital cardioplegic solution. J Thorac Cardiovasc Surg. 1993;105:995–1006.

23. Chen J, Williams S, Ho S, Loraine H, Hagan D, Whaley JM, Feder JN. Quantitative PCR tissue expression profiling of the human SGLT2 gene and related family members. Diabetes Ther. 2010;1:57–92. doi: 10.1007/s13300-010-0006-4

24. Han S, Hagan DL, Taylor JR, Xin L, Meng W, Biller SA, Wetterau JR, Washburn WN, Whaley JM. Dapagliflozin, a selective SGLT2 inhibitor, improves glucose homeostasis in normal and diabetic rats. Diabetes. 2008;57:1723–1729. doi: 10.2337/db07-1472

25. Nomura S, Sakamaki S, Hongu M, Kawanishi E, Koga Y, Sakamoto T, Yamamoto Y, Ueta K, Kimata H, Nakayama K, et al. Discovery of canagliflozin, a novel C-glucoside with thiophene ring, as sodium-dependent glucose cotransporter 2 inhibitor for the treatment of type 2 diabetes mellitus. J Med Chem. 2010;53:6355–6360. doi: 10.1021/jm100332n

26. Ishida N, Saito M, Sato S, Tezuka Y, Sanbe A, Taira E, Hirose M. Mizagliflozin, a selective SGLT1 inhibitor, improves vascular cognitive impairment in a mouse model of small vessel disease. Pharmacol Res Perspect. 2021;9:e00869. doi: 10.1002/prp2.869

27. Inoue T, Takemura M, Fushimi N, Fujimori Y, Onozato T, Kurooka T, Asari T, Takeda H, Kobayashi M, Nishibe H, et al. Mizagliflozin, a novel selective SGLT1 inhibitor, exhibits potential in the amelioration of chronic constipation. Eur J Pharmacol. 2017;806:25–31. doi: 10.1016/j.ejphar.2017.04.010

28. Vallon V, Gerasimova M, Rose MA, Masuda T, Satriano J, Mayoux E, Koepsell H, Thomson SC, Rieg T. SGLT2 inhibitor empagliflozin reduces renal growth and albuminuria in proportion to hyperglycemia and prevents glomerular hyperfiltration in diabetic Akita mice. Am J Physiol Renal Physiol. 2014;306:F194–204. doi: 10.1152/ajprenal.00520.2013

29. Stone GW, Selker HP, Thiele H, Patel MR, Udelson JE, Ohman EM, Maehara A, Eitel I, Granger CB, Jenkins PL, et al. Relationship Between Infarct Size and Outcomes Following Primary PCI: Patient-Level Analysis From 10 Randomized Trials. J Am Coll Cardiol. 2016;67:1674–1683. doi: 10.1016/j.jacc.2016.01.069

